# Persistent delirium in older hospital patients: an updated systematic review and meta-analysis

**DOI:** 10.1101/2022.01.20.22269044

**Authors:** Jonathan Whitby, Anita Nitchingham, Gideon Caplan, Daniel Davis, Alex Tsui

## Abstract

**Introduction:** Persistent delirium is recognised as a substantial problem, but there are few insights into which patient groups might be particularly affected. Delirium is associated with future dementia progression. Yet whether this occurs subclinically over months and years, or persistent delirium merges into worsened dementia is not understood.

**Methods:** We adopted an identical approach to a previous systematic review, only including studies using a recognised diagnostic framework for ascertaining delirium at follow-up (persistent delirium). We applied risk of bias assessments based on Standards of Reporting of Neurological Disorders criteria. Estimates were pooled across studies using random-effects meta-analysis, and we estimated associations with age and follow-up duration using meta-regression.

**Results:** We identified 13 new cohorts, which we added to 8 from the previous systematic review (21 relevant studies reporting persistent delirium over 37 time points). Studies were mainly at low risk of bias. Pooled delirium prevalence estimates at discharge were 40% (95% CI 24% to 54%, 10 studies). Meta-regression showed variation in prevalence of persistent delirium over time (0.6% per week, 95% CI −1.2 to −0.1, p=0.02). Older study sample age was associated with higher prevalence of persistent delirium (8.8% per SD age, 95% CI 1.1% to 17%). The rate of improvement was the same regardless of age, but the overall burden was higher with older age such that 44% (95% CI 11% to 76%) of 95-year-olds would be expected to have persistent delirium at 12 months.

**Conclusions:** This systematic review emphasises the key importance of delirium as a persistent and extensive problem, particularly in the oldest-old. Addressing persistent delirium will require a whole-system, integrated approach in order to detect, follow-up and implement opportunities for recovery across all healthcare settings.

## Introduction

Delirium is widely recognised as having a substantial impact on healthcare for older people, with several clinical guidelines serving to address its detection, prevention and management in hospitals.^4, 5^ However, many questions remain over the natural history of delirium, particularly its relationship with dementia.^6^ For example, it is not clear if underlying dementia leads to a more severe or prolonged delirium.^7^ We know that delirium is associated with future dementia progression.^1-3^ Yet whether this occurs subclinically over months and years, or persistent delirium merges into worsened dementia is not understood.

Persistent delirium has been of interest since a systematic review from 2009 suggested one in five cases were still evident six months after discharge.^8^ Subsequently, the number of older people presenting for urgent and emergency care has increased.^9^ There has also been a consistent trend for more acute presentations of people living with dementia; around half are admitted within the first 12 months of diagnosis.^10^ In light of these changes, we set out to update the systematic review to provide current estimates for the epidemiology of persistent delirium.

## Methods

### Eligibility criteria

We followed the 2020 PRISMA guidance.^11^ We used the same criteria applied in the previous systematic review: (i) study population of at least 20 hospital patients; (ii) patients aged ≥50 years; (iii) prospective study with follow-up of at least one week; (iv) acceptable definition of delirium at enrolment. We excluded studies investigating delirium in critical care and in the context of terminal illness or palliative care. Our only modification was to require studies to use a recognised diagnostic framework for ascertaining delirium at follow-up (persistent delirium); studies from the original systematic review meeting this criterion were carried over into the current analysis. Given this was an update of a previous systematic review, we did not devise a *de novo* protocol for PROSPERO.

### Outcome measures

The primary outcome was proportion of patients with delirium at follow-up, where the denominator was the number of participants who had delirium at inception. We considered any definition based on the Diagnostic and Statistical Manual (DSM), International Classification of Disease, the Confusion Assessment Method (CAM) or the Delirium Index to be sufficiently detailed to ascertain delirium reliably.

### Search strategy

Updating the original review, we searched from 1 year before the previous end date (September 2006) to 11^th^ January 2022. We searched the same electronic databases: MEDLINE, EMBASE, PsycINFO and the Cochrane Database of Systematic Reviews, using the following search terms: Delirium [Title] AND (prognosis OR outcome OR aged OR occurrence) [Title/Abstract], replicating the original search strategy. We confirmed the sensitivity of the search by ensuring that we captured all studies from the previous review.

### Data collection and study selection

Covidence (www.covidence.org, Veritas Health Innovation Ltd.) was used to manage the abstract and full-text screening, assessing risk of bias and data extraction. Three researchers independently reviewed titles and abstracts (J.W., A.N., A.T.) to determine the eligibility. Conflicts were resolved by discussion and consensus. The same reviewers extracted data using a *pro forma*.

### Assessment of quality and biases

We determined risk of bias based on the framework outlined by the Standards of Reporting of Neurological Disorders (STROND) criteria.^12, 13^ We considered bias arising (i) from specific patient settings, e.g., general medical patients, cohort with intracerebral haemorrhage; (ii) from sample selection, e.g., convenience, consecutive; (iii) from sample criteria, e.g., excluding patients unable to consent, assessed as being too sick; (iv) from reference standard used, e.g., DSM, CAM; (v) from expertise in applying the reference standard, e.g. routine data, dedicated researcher. We rated studies high, low or unclear risk of bias in each of the five domains. Quality of evidence was assessed using the GRADE framework, which considers risk of bias, inconsistency, indirectness, imprecision and publication bias.

### Statistical analysis

We extracted summary statistics for the prevalence of persistent delirium at each time point and calculated their standard errors (sqrt [p (1-p) / n)]. We assumed methodological heterogeneity across studies, accounting for this using DerSimonian–Laird random-effects models.^14^ Statistical heterogeneity was assessed with the I^2^ statistic. Meta-regression was used to estimate the relationship between persistent delirium and follow-up time, mean age, and dementia prevalence in each study sample. We plotted marginal predictions for any parameter associated with persistent delirium. To assess publication bias, we plotted the estimated proportion of delirium occurrence against the standard error of that estimate and inspecting the degree of asymmetry. We used Stata 16.1 (StataCorp, Texas) for all analyses.

## Results

We identified 6474 articles, screening 5556 after removal of duplicates (see PRISMA diagram). We assessed 115 full-text articles for eligibility. We identified 13 new cohorts, which we added to eight from the previous systematic review, giving a total of 21 relevant studies reporting persistent delirium over 37 time points (Table 1).^15-34^ Most studies were in medical patients and they were all from high-income settings and casemix by ethnicity was not generally reported. Samples ranged in size (n=23 to n=278) and duration of follow-up (up to 12 months). Average age and proportion with dementia were reported as 82.3 years and 43% respectively, though both varied substantially between different study cohorts.

**Table 1.** Characteristics of included studies.

| Study | Setting | N | Age<br>(years) | Dementia<br>(%) | Delirium<br>ascertainment | Discharge | 2 weeks | 1 month | 3 months | 6 months | 12 months |
| --- | --- | --- | --- | --- | --- | --- | --- | --- | --- | --- | --- |
| Levkoff 1992 | M, S | 91 | 81 |  | DSM-III | 58% |  |  | 37% | 31% |  |
| Rudberg 1997 | M, S | 64 | 75 | 33 | DSM-III-R | 11% |  |  |  |  |  |
| Marcantonio<br>2000 | S | 52 | 79 | 66 | CAM | 38% |  | 29% |  | 6% |  |
| Kelly 2001 | M | 61 | 89 |  | CAM | 72% |  | 46% | 25% |  |  |
| McCusker 2003 | M | 181 | 83 | 70 | DI | 32% |  |  |  | 32% | 41% |
| Kiely 2004 | PAC | 85 | 85 | 23 | CAM |  |  | 51% |  |  |  |
| McAvay 2006 | M | 55 | 80 | 27 | CAM | 24% |  |  |  |  |  |
| Inouye 2007 | M | 169 | 80 | 20 | CAM | 51% |  |  |  |  |  |
| Lundstrom 2007 | S | 129 | 82 | 32 | DSM4 | 12% |  |  |  |  |  |
| Kiely et al. 2009 | PAC | 412 | 84 | 38 | CAM |  | 67% | 56% | 40% | 32% |  |
| McManus 2009 | Stroke | 23 | 75 |  | CAM |  |  | 64% |  |  |  |
| Arinzon 2011 | M | 92 | 80 | 90 | CAM, DRS |  | 50% |  |  |  |  |
| Lee 2011 | S | 70 | 80 | 10 | CAM |  |  | 20% |  | 3% | 3% |
| Witlox 2013 | S | 27 | 84 |  | CAM |  |  |  | 19% |  |  |
| Velilla 2013 | M | 45 | 87 |  | CAM, DSM-IV |  |  | 18% |  |  |  |
| Cole 2015 | M, S | 278 | 85 | 55 | CAM |  |  | 73% | 61% |  |  |
| Vasunilashorn<br>2016 | M, S | 250 | 79 |  | CAM | 24% |  |  |  |  |  |
| Miu 2016 | PAC | 89 | 84 |  | CAM | 79% |  | 85% | 55% |  |  |
| Jackson 2016 | M | 125 | 84 | 36 | DSM-IV |  |  |  | 6% |  |  |
| Cole 2017 | M, S | 152 | 85 | 52 | CAM |  |  | 63% |  |  |  |
| Reznik 2021 | Stroke | 590 | 71 | 13 | DSM-5 | 48% |  |  |  |  |  |

% reported refer to the denominator of patients with delirium in the original sample.
M medical; S surgical; PAC post-acute care; DSM Diagnostic and Statistical Manual; CAM Confusion Assessment Method; DI Delirium Index

In the main, studies were at low risk of bias insofar as most followed a sample representative of the target setting (e.g., acute hospital care) and used consistent outcome ascertainment instruments (GRADE certainty rating for risk of bias = ‘low’) (Figure 2). Around half excluded patients unable to give consent (and did not report procedures to allow proxy consent) and/or excluded participants too sick to assess for delirium. Case ascertainment was based on a consensus definition (e.g. DSM) in 6 studies; the rest used instruments such as CAM. Given each has different degrees of restrictiveness in their definition,^35^ the GRADE certainty rating for indirectness was ‘moderate’.

**Figure 1.**
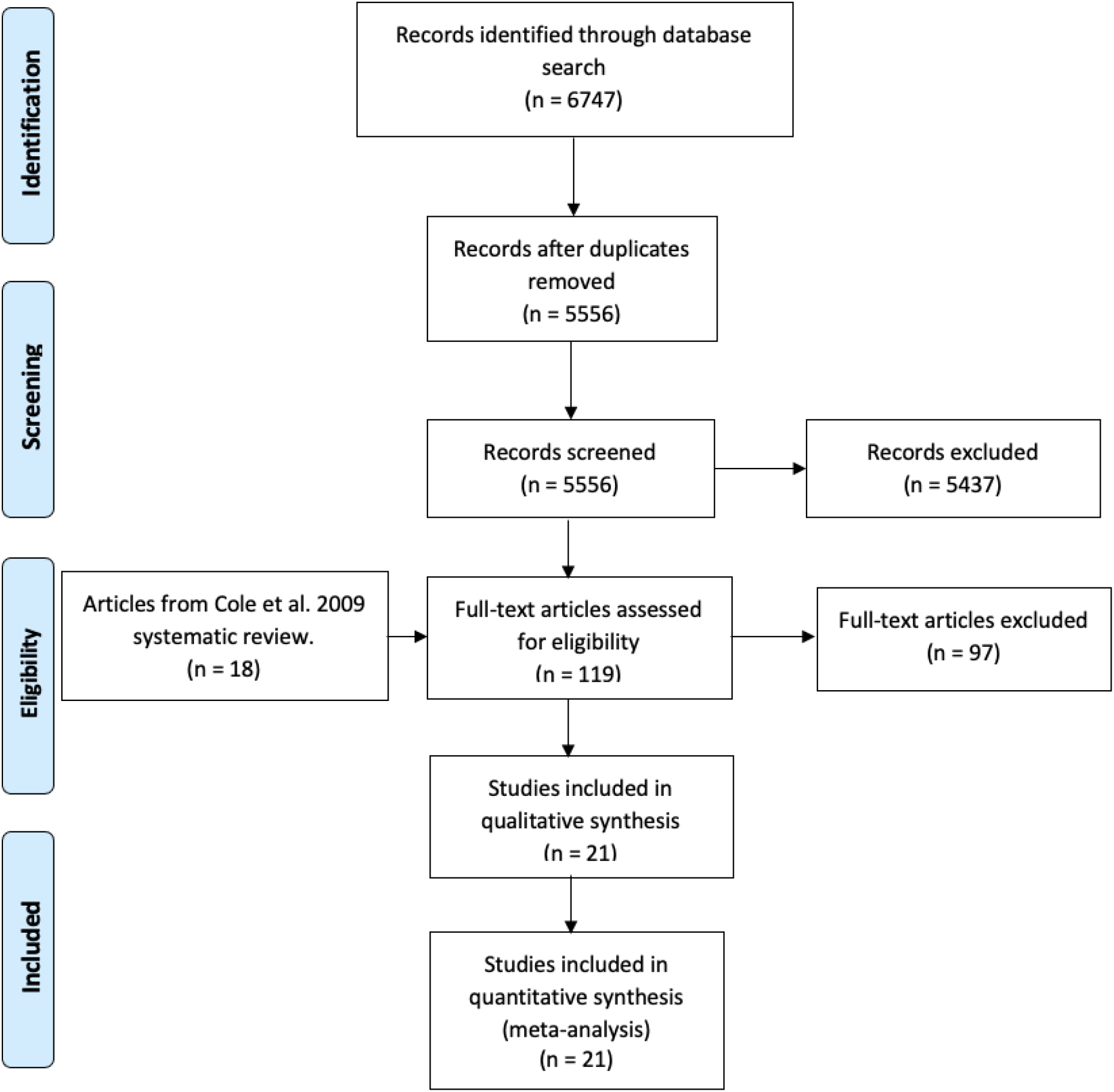
PRISMA flow detailing study selection process.

**Figure 2.**
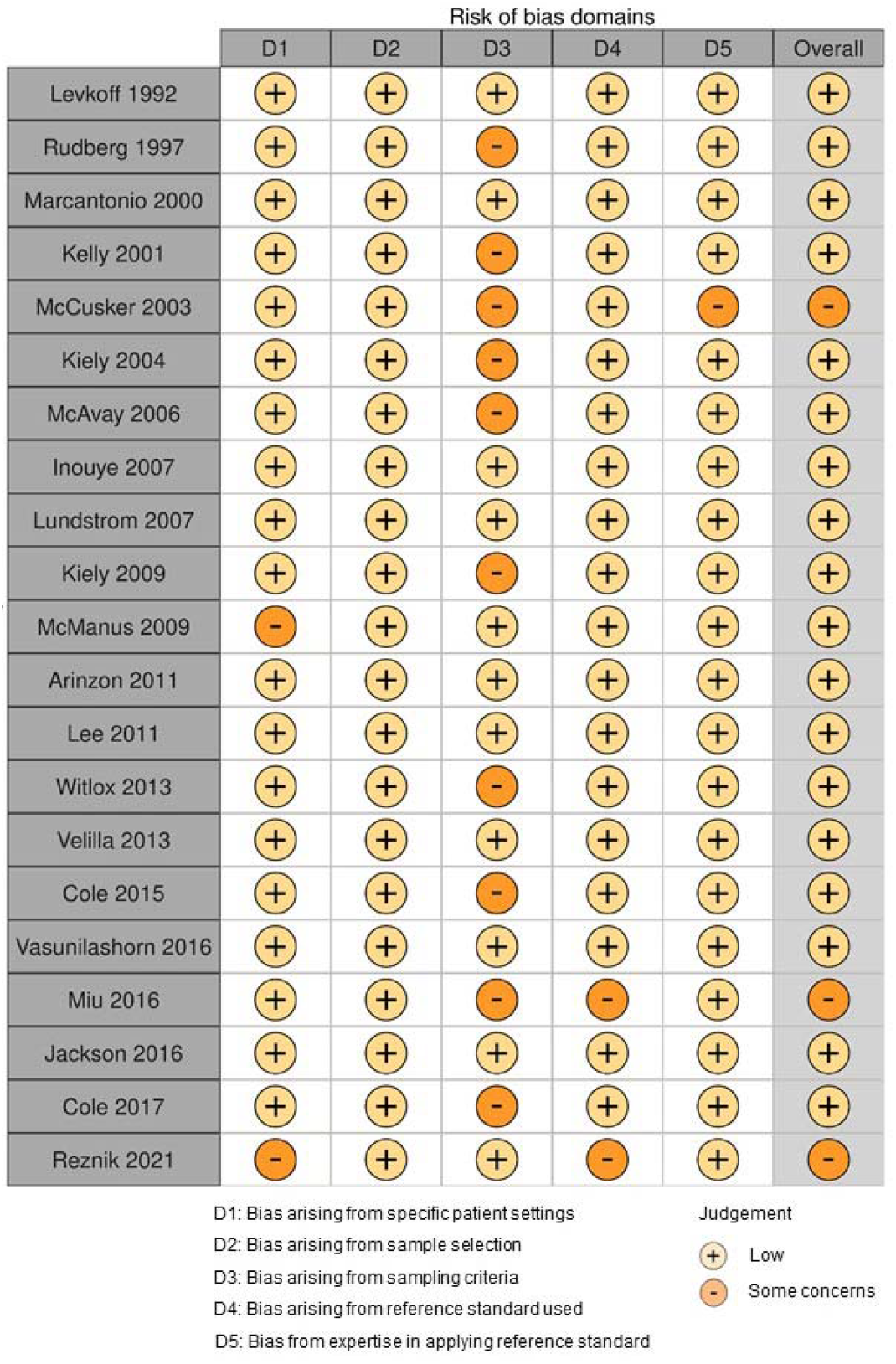
Assessment of risk of bias.

**Figure 3.**
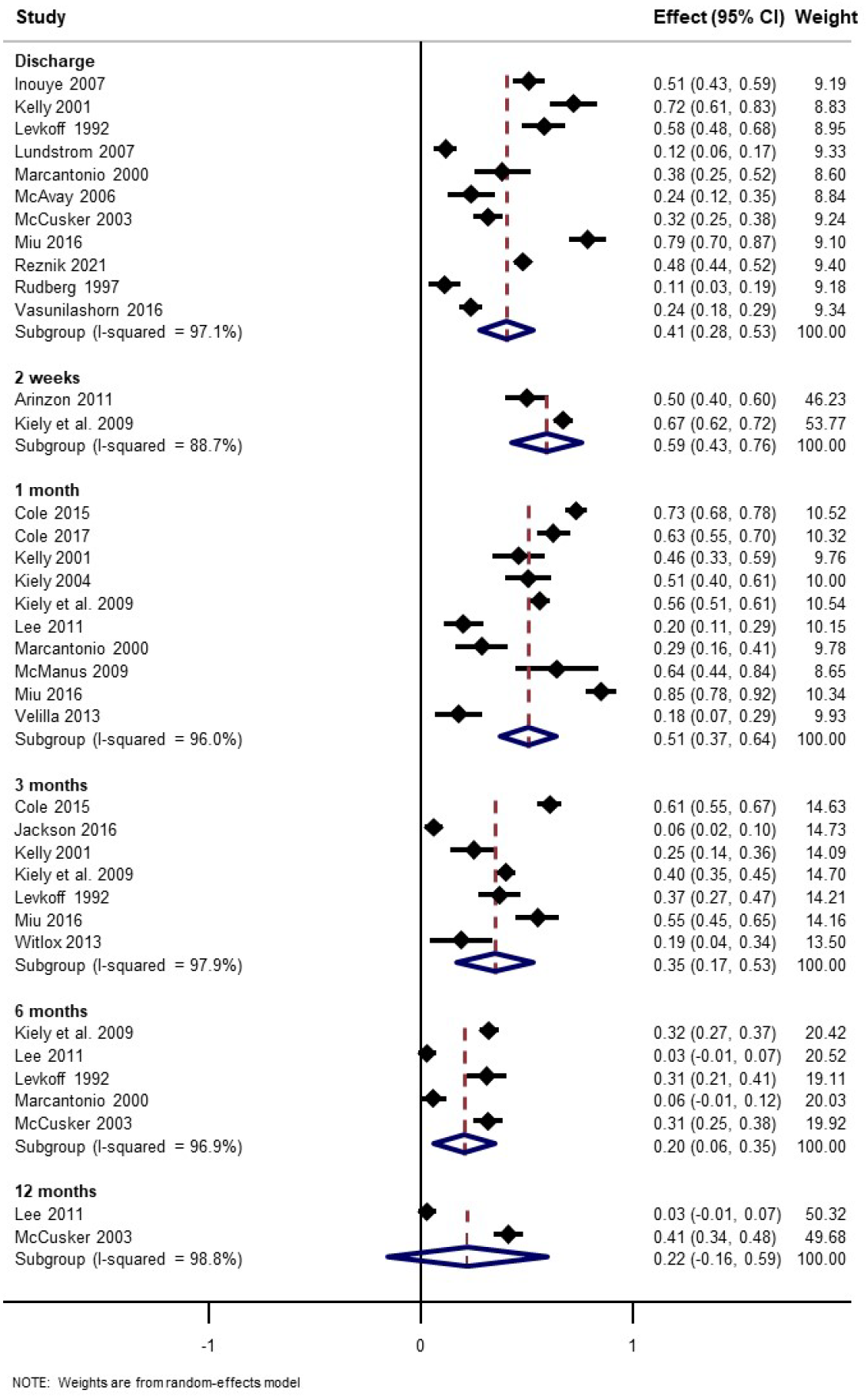
Forest plot showing pooled estimates of persistent delirium over time.

Pooled delirium prevalence estimates at discharge were 40% (95% CI 24% to 54%, 10 studies) (Figure 4). Although there were few studies with data beyond 6 months, each time point reported considerable persistent cases (pooled estimates ranging from 20% to 59%). There was substantial statistical heterogeneity at all time points (all subgroup I^2^ >89%) (GRADE certainty rating for imprecision = moderate). Funnel plots of prevalence versus the standard error of the estimate did not demonstrate any asymmetry, leading to a GRADE certainty rating for publication bias of ‘high’.

**Figure 4.**
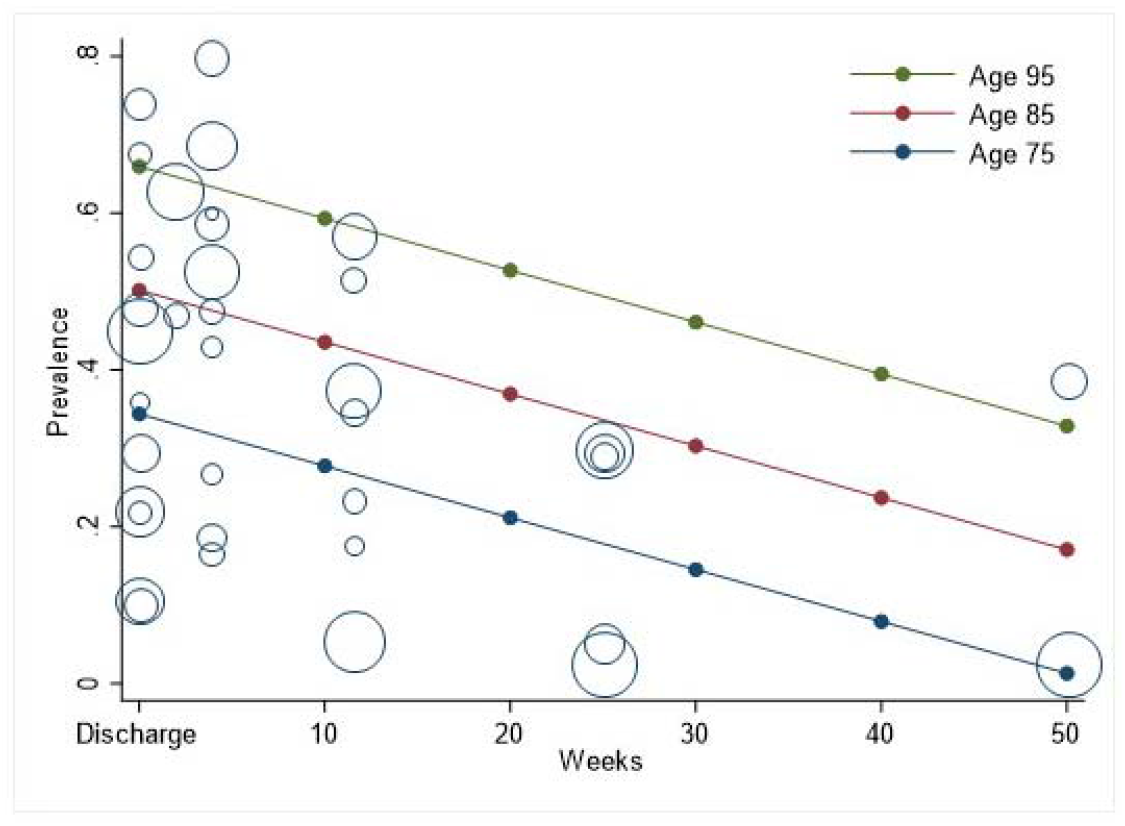
Meta-regression showing estimated prevalence over time, with marginal predictions stratified by age.

Meta-regression showed variation in prevalence of persistent delirium over time (0.6% per week, 95% CI −1.2 to −0.1, p=0.02) (Table 2). This monotonic decrease in prevalence over time led to a GRADE certainty rating for inconsistency as ‘low’. Older study sample age was associated with higher prevalence of persistent delirium (8.8% per SD age, 95% CI 1.1% to 17%). Persistent delirium did not vary with the proportion of study sample with dementia, though only 14 studies reported this. Figure 4 shows the predicted meta-regression margins for three age groups over time. The rate of improvement was the same regardless of age, but the overall burden was higher with older age such that 44% (95% CI 11% to 76%) of 95-year-olds would be expected to have persistent delirium at 12 months. The overall GRADE certainty for this estimate is ‘moderate’.

**Table 2.** Meta-regression estimating the proportion of individuals with persistent delirium

|  | Univariable |  |  |  | Multivariable |  |  |  |
| --- | --- | --- | --- | --- | --- | --- | --- | --- |
| | $\beta$ | 95% CI | | p | $\beta$ | 95% CI | | p |
| Time (weeks) (n=37) | -0.7 | -1.2 | -0.1 | 0.02 | -0.6 | -1.2 | -0.1 | 0.02 |
| Age (per SD) (n=37) | 9.1 | 0.9 | 17.4 | 0.03 | 8.8 | 1.1 | 16.5 | 0.03 |
| Dementia % (n=24) | 0.3 | -0.1 | 0.8 | 0.14 |  |  |  |  |
| $\beta$ represents % change in prevalence of persistent delirium. Dementia refers to % with dementia in the study sample | | | | | | | | |

## Discussion

We showed that persistent delirium remains a substantial problem well beyond the acute phase. This persistence does not appear to vary with baseline dementia, albeit using a study-level ecological measure of dementia. A clearer association was evident with increasing age. Taken together, these findings suggest a considerable need to focus delirium recovery for both individuals and health services.

Our data should be considered in light of a number of limitations. Other than duration, the variables we could use for meta-regression relied on study-level data on mean sample age and dementia prevalence. This limits the interpretation due to residual confounding for quantities such as frailty. Most studies had significant attrition due to mortality and loss to follow up, though this might be expected to underestimate the true prevalence of persistent delirium. All studies were performed in high-income countries, making it difficult to generalise our findings to other settings. Many studies only included delirium detected in the first 24-48 hours of admission, which could have led to under-ascertainment. There is likely to be a degree of overlap between the reports of *delirium at discharge* and some of the earlier follow-up time points (*2-weeks* and *4-weeks*) because the former category encompassed a variable duration. Some publications only reported persistent delirium at discharge, so it is impossible to know how long these patients remained as cases. Similarly, it is unclear how long delirium might have been present prior to transfer for individuals admitted to post-acute care. We also restricted our search to studies published in English. Finally, we detected significant heterogeneity between studies, though we attempted to limit this by defining our outcome measure as precisely as possible.

In updating the original systematic review with 15 years’ worth of new studies, we have extended those findings to demonstrate the highest prevalence of persistent delirium with older age. While older age is likely to be a proxy for dementia and frailty, the gradient suggests that delirium may never resolve in the oldest-old. More broadly, these data are consistent with terminal decline in cognition observed longitudinal studies.^36^ Higher education is associated with delayed onset, though a faster trajectory once terminal decline begins.^37^ Though previous cohorts have not considered delirium to be a driver, the Delirium and Population Health Informatics Cohort has that shown the largest post-delirium decline in cognition occurs in those with *higher* baseline cognition.^38^ Dedicated prospective studies are needed to fully capture the influence of evolving dementia reciprocally affecting persistent delirium.

The clinical implications of our findings indicate an urgent need to develop delirium recovery and cognitive rehabilitation services. These are almost non-existent, certainly out of proportion with the potential demand, though multicomponent interventions show promise.^39^ Recognising the opportunities for the emerging field of interface acute geriatrics would be an important starting point,^40^ and continued community-based management of delirium is likely to have an impact.^41^

This systematic review emphasises the key importance of delirium as a persistent and extensive problem, particularly in the oldest-old. Addressing persistent delirium will require a whole-system, integrated approach in order to detect, follow-up and implement opportunities for recovery across all healthcare settings.

## Data Availability

Data can be shared on request

## Acknowledgements

JW is funded through a grant from the Dunhill Medical Association, DD is funded by the Wellcome Trust (WT107467). AT is an Alzheimer’s Society Clinical Research Training Fellow. The MRC Unit for Lifelong Health and Ageing at UCL received core funding through the Medical Research Council (MC_UU_00019/1).

